# Estimating optimal therapeutic drug levels of anti-tuberculosis medications based on treatment safety and effectiveness

**DOI:** 10.1101/2024.08.30.24312723

**Authors:** Gustavo Amorim, David W. Haas, Marcelo Cordeiro-Santos, Afrânio L. Kritski, Marina C. Figueiredo, Cody Staats, Brian Hachey, Megan Turner, Bruno B. Andrade, Valeria C. Rolla, Timothy R. Sterling, the Regional Prospective Observational Research in Tuberculosis (RePORT)-Brazil network

**Affiliations:** Vanderbilt University Medical Center, Department of Biostatistics, Nashville, TN; Vanderbilt University Medical Center, Department of Medicine, Division of Infectious Diseases, Nashville, TN; Meharry Medical College, Department of Internal Medicine, Nashville, TN; Fundação Medicina Tropical Dr. Heitor Vieira Dourado, Manaus, Brazil; Universidade do Estado do Amazonas (UEA), Manaus, Brazil; Universidade Federal do Rio de Janeiro, Faculdade de Medicina, Rio de Janeiro, Brazil; Multinational Organization Network Sponsoring Translational and Epidemiological Research (MONSTER) Initiative, Bahia, Brazil; Instituto Brasileiro para Investigação da Tuberculose, Fundação José Silveira, Salvador, Bahia, Brazil; Instituto Gonçalo Moniz, Fundação Oswaldo Cruz, Salvador, Bahia, Brazil; Universidade Salvador (UNIFACS), Laureate Universities, Salvador, Bahia, Brazil; Escola Bahiana de Medicina e Saúde Pública (EBMSP), Salvador, Bahia, Brazil; Faculdade de Tecnologia e Ciências (FTC), Salvador, Bahia, Brazil; Instituto Nacional de Infectologia Evandro Chagas, Fundação Oswaldo Cruz, Rio de Janeiro, Brazil

**Keywords:** tuberculosis, pharmacokinetics, safety, effectiveness, drug monitoring

## Abstract

**Background:** Therapeutic drug ranges (TDR) for standard anti-tuberculosis (TB) treatment have been determined based on expected drug levels at least 2 hours after taking the dose. In this study we constructed TDR for TB drug levels based on minimizing drug toxicity and maximizing treatment effectiveness.

**Methods:** Participants were followed prospectively in the Regional Prospective Observational Research in Tuberculosis (RePORT)-Brazil observational cohort study. We focused on participants with culture-confirmed drug-susceptible pulmonary TB who underwent standard TB therapy. TDR were estimated for each TB drug separately: isoniazid (INH), rifampin (RIF), ethambutol (EMB), and pyrazinamide (PZA). TDR were defined as drug concentrations that were both safe and effective: safety was defined as the probability of having an ADR of at most 5%, while effectiveness was defined as a probability of at least 95% of not having either TB treatment failure or recurrence.

**Results:** There were 765 plasma samples from 448 patients; 110 (24.6%) were people with HIV, 9 (2.0%) had a grade 3 or higher ADR, and 15 (3.3%) had treatment failure/recurrence. Higher drug concentrations of INH, RIF and EMB were associated with increased odds of having ADR. High concentrations of INH suggested protection against treatment failure/recurrence. Estimated therapeutic drug range for INH (2.3-8.2 µg/ml) and for RIF (0.5-7.5 µg/ml) differed from the currently recommended drug ranges (3-5 µg/ml and 8-24 µg/ml, respectively). Estimates for PZA and EMB were similar to the currently recommended values.

**Conclusions:** Our estimated upper end TDR were higher for INH and lower for RIF compared to currently recommended ranges.

## Introduction

Tuberculosis (TB) is a major global health threat worldwide. Over 7.5 million people were diagnosed with TB in 2022 and 1.3 million died in the same year. [1] Standard anti-tuberculosis (TB) therapy includes two months of isoniazid (INH), rifampin (RIF), pyrazinamide (PZA), and ethambutol (EMB), followed by four months of isoniazid and rifampin. [1, 2]

TB treatment is highly effective in most patients, with cure rates as high as 97% among those who adhere to treatment in randomized controlled trials.[3] However, in real-world settings, approximately 88% of TB patients have a successful treatment outcome.[1] Many patients experience drug toxicity, acquire drug resistance during therapy, fail the standard treatment regimen, are lost to follow-up, or stop taking treatment.[4]

Treatment failure occurs in fewer than 5% of treated TB patients. TB recurrence, in which individuals successfully complete treatment but later have signs and symptoms of active TB, can be due to relapse or exogenous reinfection with another strain of *M. tuberculosis*. Approximately 4% of patients treated with standard TB therapy experience relapse or require retreatment within two years.[5, 6]

Treatment outcomes such as safety and effectiveness have been shown to be associated with levels of at least some of the standard anti-tuberculosis medications. [7–11] However, the therapeutic ranges for standard anti-TB treatment have been determined based on expected levels two or six hours (the latter if there is delayed absorption) after taking the dose [12, 13]. In this study we took a different approach and constructed therapeutic ranges for TB drug levels based on concomitantly minimizing drug toxicity and maximizing treatment effectiveness. It is also important that such evaluations include all of the TB drugs given in combination, since these drugs are given together in the clinical setting, and the drug combination affects both toxicity and effectiveness.

We performed an observational cohort study in which persons with drug-susceptible culture-confirmed pulmonary TB were treated with standard therapy and followed prospectively for treatment toxicity and effectiveness. We evaluated the association between plasma TB drug exposure and toxicity and effectiveness outcomes and estimated therapeutic drug ranges for which concentrations were safe and effective.

## Methods

### Study design and population

Our study population consisted of participants enrolled in Regional Prospective Observational Research in Tuberculosis (RePORT)-Brazil, a prospective observational cohort of individuals with newly-diagnosed, culture-confirmed, pulmonary TB.[14] Patients were enrolled from five locations in Brazil, from 2015 to 2019. Participants were followed for 24 months from enrollment. Our analyses were limited to persons who had TB susceptible to INH and RIF and received standard anti-TB treatment with these medications. All study participants provided written informed consent. The RePORT-Brazil study protocol was approved by the institutional review boards at all study sites in Brazil and at Vanderbilt University Medical Center.

### Data collection and definitions

Demographic and clinical data were collected for each study participant throughout the study. Blood, urine, and plasma samples were collected at all four in-person study visits: at baseline (initiation of TB treatment), months one and two after treatment initiation, and at treatment completion, typically 6 months after baseline. Follow-up visits were performed by telephone every six months after treatment completion; persons with symptoms of TB were evaluated in person. All data were entered and managed using REDCap (Research Electronic Data Capture) [15, 16]. All participants underwent HIV testing at baseline unless already diagnosed with HIV.

Plasma drug concentrations for INH, RIF, PZA, EMB, and desacetyl-rifampin (des-RIF) were quantified by liquid chromatography-mass spectrometry (LC/MS-MS) with a SCIEX 6500 Qtrap mass spectrometer (Redwood City, CA.). Stored plasma samples were collected within 24 hours of the previous TB drug dose at up to four time points (baseline, months 1, 2, and end of treatment) for each participant and frozen at -80 ºC. We focused on time points at which drug levels should have been present in plasma, and which could have predicted subsequent toxicity or effectiveness. Therefore, we restricted our analysis to patients with plasma collected within 8 hours of last TB drug medication during the intensive phase of TB therapy (first two months). Plasma concentrations below the level of quantification (BLQ) were set to half of the BLQ cut-off values. BLQ cut-off values were 100 ng/mL for INH and RIF, 500 ng/mL for PZA, and 50 ng/mL for EMB. Drug concentrations were determined after TB treatment completion, thus were not available to modify drug dosing during treatment.

Outcomes of interest included TB treatment effectiveness (i.e., treatment failure and recurrence) and toxicity (adverse drug reactions (ADR) grade 3 or higher that were related to TB treatment). Treatment failure was defined as remaining sputum culture-positive or smear-positive at month 5 or later during treatment. Recurrence was defined as culture-confirmed TB or symptoms consistent with TB after the participant had been cured or completed TB treatment.

The therapeutic drug range was defined as drug concentrations that were both safe and effective. We defined safety as the probability of having an ADR of at most 5%, while efficacy was defined as a probability of at least 95% of not having either treatment failure or TB recurrence. Therapeutic ranges were estimated for each drug separately: INH, RIF, PZA, and EMB.

### Statistical analysis

Our goal was to compute a therapeutic range in which the concentration of the four standard TB drugs were both safe and effective. We used a 2-step approach to construct therapeutic ranges. In the first step we related patients’ drug concentrations (collected within 8 hours since last dose) to toxicity and effectiveness outcomes via regression models. Next, using pre-defined bounds for probabilities of having ADR and treatment failure/recurrence, we estimated a range for drug concentrations collected at 2 hours after taking the most recent dose that was both safe and effective.

Relating drug concentrations to toxicity and effectiveness outcomes, however, is challenging. A high or low individual drug concentration is intrinsically connected to when the medication was taken; drug concentration reaches a maximum at 2-3 hours after taking the medicine and decreases non-linearly over time. To overcome this issue, we took two approaches to estimate the therapeutic range in a meaningful and interpretable way: 1) we created individual drug percentiles that resembled the well-known z-score in growth curves and 2) we used machine learning methods to predict maximum drug concentrations (Cmax) at the patient level.

Patient-percentiles were estimated from a quantile regression model in which the observed drug concentration was regressed against the timing of the most recent TB dose. We used restricted cubic splines with three knots placed in the 1^st^, 2^nd^, and 3^rd^ quartiles, and modelled drug concentration on the natural log scale. This approach is similar to that of Jamsen and colleagues [17], as an alternative to discrete time intervals to estimate individual drug exposures. The expected median drug concentration, given by the fitted quantile regression, was compared to the observed drug concentration to derive standardized scores (z-scores) that ranged from 0 to 1. High (close to 1) and low (close to 0) scores represented observations far above or far below the expected median concentration, respectively. Technical details about derivation of individual percentiles of drug exposures are in **Appendix A**. With the patient-percentile computed for each patient, we used generalized estimating equations (GEE) with the logit link to regress the outcomes on the (logit of the) patient-percentiles, and then estimated the probability of having an ADR or failure/recurrence. We used patients as clusters and assumed an independent working correlation matrix. Once the model was fitted, we calculated the percentile associated with a probability of having ADR equal to 5%. This percentile, say *pp*_*ADR*=5%_, represents an upper safety limit (safety bound); patients with higher percentiles than *pp*_*ADR*=5%_ have higher odds of having adverse events. Next, to make the bound interpretable, we back-transformed it into the original concentration scale (ug/ml) and then evaluated the expected concentration that is safe at 2 hours after the most recent dose. The effectiveness bound, the minimum drug concentration that is effective, was calculated similarly. We used GEE to regress the chance of having failure/recurrence on patient percentiles. Then, we computed the percentile associated with a probability of failure/recurrence equal to 5%. This is the effectiveness bound. Again, to make this bound interpretable, we back-transformed it into the original concentration scale and computed the effectiveness bound at 2 hours after taking the last dose. We repeated the procedure above for both upper (safety) and lower (effectiveness) bounds for the drug concentration for 999 bootstrap replicates. This allowed us to calculate 95% confidence intervals (CI) for our safety and effectiveness bounds. We used the bootstrap percentile method to calculate the 95% CI.

In addition to using the patient-percentile to compute therapeutic drug ranges, we also used extreme gradient boosting (XGBoost) to create therapeutic drug ranges using the predicted Cmax. We used a 10-fold cross-validation to predict the Cmax at the patient-level using the following baseline covariates: age, sex, body mass index (BMI), HIV status, smoking and alcohol status, illicit drug use, hemoglobin A1C (HbA1c), ancestry informative markers, whether a patient was assigned directly observed therapy (DOT) at baseline or not, as well as *NAT2* acetylator status (when modelling INH), and time since most recent dose. The predicted Cmax was correlated to the probability of ADR and treatment failure/recurrence, also using GEE with the logit link and independent correlation matrix. The maximum drug concentration that was safe and effective at 2 hours after taking the last dose was estimated from the fitted models. A total of 999 bootstrapping replicates were used to obtain 95% CIs.

All analyses were performed using the statistical software R, version 4.3.1. Three patients were excluded because they had drug concentrations BLQ for all drugs, at all visits. For association analyses between drug levels and outcomes, described below, we used data from baseline, month 1, and month 2, except if a patient had an adverse event; then, only data up to the adverse event date were used in the analysis.

## Results

This study included 448 RePORT-Brazil participants who had at least one plasma sample collected within 8 hours of their most recent TB drug dose at either baseline, month 1, or month 2; the median age was 35 years old (inter-quartile range [IQR]: 25 47), most were male (67.6%), and 110 (24.6%) were people living with HIV (PWH). About half of the patients were slow *NAT2* acetylators. Overall, 9 (2.0%) participants had a grade 3 or higher ADR and 15 (3.3%) had treatment failure or TB recurrence (**Table 1**). Among the 448 participants, 765 plasma samples were assayed and included in this study: 113 (14.8%) at baseline, 332 (43.4%) at month 1, and 320 (41.8%) at month 2. The distribution of each drug concentration in relation to the first eight hours since last dose is displayed in **Figure 1**. The red bars correspond to the currently recommended therapeutic ranges: 3-5 µg/ml for INH, 8-24 µg/ml for RIF, 2-6 µg/ml for EMB, and 20-50 µg/ml for PZA [12, 13]. For many participants, plasma drug concentrations were below the currently recommended therapeutic ranges for RIF, PZA, and EMB, while the majority of patients had higher drug concentrations than the recommended range for INH. Such variations, and in particular the high proportion of low RIF concentrations, compared to recommended ranges have also been observed elsewhere. [18]

Patient-percentiles were associated with grade 3 or higher ADR for all four drugs (**Table 2**). High patient-percentiles, (i.e. concentrations above the median drug concentration observed across the study population) derived from INH and RIF were, in particular, strongly associated with increased log-odds of treatment-related adverse event: Log-odds = 0.53 (95%CI = [0.29; 0.77]) and 0.76 (95%CI = [0.25; 1.27]), respectively. Similar patterns were observed when regressing ADR on (log-transformed) Cmax. High Cmax values for INH, RIF, EMB were associated with higher odds of ADR: Log-odds = 5.18 (95% CI = [2.26; 8.10]), 4.38 (95% CI = [1.05; 7.71]), and 5.20 (95% CI = [0.16; 10.24]), respectively. Although not statistically significant at the 5% level, high Cmax for PZA also suggested a positive correlation with higher odds of ADR.

Associations between the exposures of interest, patient-percentile or Cmax, with treatment failure/recurrence were modest (**Table 3**). High Cmax for INH was protective against failure/recurrence, with a log-odds of -0.16 (95% CI = [-0.29; -0.02]). A similar pattern was observed for patients with high percentile (patient-percentile approach, for INH): higher values were protective against failure/recurrence (log-odds = - 0.70, 95% CI = [-1.22; -0.17]). Analyses for the other TB drugs did not reach statistical significance at the 5% level.

The estimated safety and effectiveness bounds, computed at 2 hours after taking the medicine, for both patient-percentile and Cmax approaches, are displayed in **Table 4**. Overall, the results were comparable between the two approaches. Due to the moderate/strong associations between the exposures (patient-percentile or Cmax) and the ADR, safety bounds could be computed with some accuracy. However, as there was weak/no association with treatment failure/recurrence, the effectiveness bound could not be computed for most drugs and approaches.

## Discussion

In this study we estimated a therapeutic drug range that is safe and effective, for all four main TB drugs, conducted within a prospective cohort of TB patients in Brazil. Concentrations of INH, RIF, EMB, and PZA varied substantially and, despite many individuals having drug concentrations lower than target therapeutic ranges, the overall risks of unfavorable effectiveness and toxicity outcomes were low.

Our estimation was based on pre-defined levels of probabilities for drug toxicity and TB treatment failure or recurrence. In other words, we were interested in estimating ranges in which the drugs were both safe and effective. We approached this problem via two routes: 1) using patient-percentiles to summarize drug concentrations relative to the median drug concentration observed in the population, and 2) using machine learning methods to model the non-linear effect of drug concentration over time and extract the maximum drug concentration Cmax.

Although both patient-percentile and Cmax approaches showed largely similar results, they seemed to differ from therapeutic ranges reported in the literature, in particular for INH. Here, using Cmax, our estimated range of 2.31-8.22 (95% CI = [0; 3.88] for the lower bound and 95% CI = [7.50; 11.00] for the upper bound) was higher than the 3-5 µg/ml range recommended by [13]. That is, in our setting, patients with INH drug concentrations (Cmax) lower than 8.22 (95% CI = [7.50; 11.00]) had low chances (less than 5%) of having adverse drug reactions. Estimates for the upper bound of the therapeutic ranges, i.e., levels associated with safety, for both EMB and PZA fell into the currently recommended ranges. For RIF, our estimated safety bound was 7.49 [6.68; 9.71], closer to the lower end of the 8-24 µg/ml recommended range.

We also noticed a positive association between drug concentrations and increased odds of having grade 3 or higher ADR. Similar results have been observed elsewhere, in particular for INH and RIF. [19, 20] At the same time, higher levels of INH were protective against TB treatment failure/recurrence. No other drugs were associated with treatment failure/recurrence, at the 5% level. This affected the calculation of the effectiveness bound, in particular for the patient-percentile approach, as finding the exposure that would result in a 5% chance of having treatment failure/recurrence would require extrapolating outside the (0,1) interval.

This study had several limitations. First, data were collected sparsely, with only one observation collected from each patient per visit. The number of outcomes was lower than anticipated; and due to the limited number of events, we were unable to adjust for covariates or interactions with other drugs levels in our models, because of the risk of overfitting. We were also unable to fit more complex models, for example using non-linear models to alleviate some linearity assumptions between exposures (Cmax or patient-percentiles) and outcomes of interest. In addition, for most participants, we only had aggregated monthly DOT data, so we had to rely on self-reported timing of previous drug doses and were not able to take into account how adherence could affect our results. Finally, it is important to highlight once more that our estimated therapeutic ranges were based on pre-defined probabilities of having ADR or TB treatment failure/recurrence. Different probabilities would lead to different therapeutic ranges.

With the above limitations noted, we believe that our findings complement existing research and provide new insights for future work for TB therapeutic drug monitoring (TDM). According to the suggestions by Verbeek et al [21], to justify TDM, research must demonstrate: 1) a clear relationship between drug plasma concentration and therapeutic response; 2) a known and narrow therapeutic window for measuring drug plasma concentration; 3) the inability to individualize dosage on clinical indicators alone; and 4) a large variability in interindividual pharmacokinetic variability.[4] With these goals in mind, future work should aim to confirm the ideal range of drug concentration, perhaps especially for INH, that minimizes toxicity and maximizes effectiveness.

## Data Availability

All data produced in the present study are available upon reasonable request to the authors

## Acknowledgments

The RePORT-Brazil network included the following study sites and investigators:

**Instituto Nacional de Infectologia Evandro Chagas, Fiocruz, Rio de Janeiro, Brazil:** Valeria Rolla, Maria Cristina Lourenço, Aline Benjamin, Quezia Medeiros, Felipe Ridolfi, Adriano Gomes, Marcus Rockembach

**Secretaria Municipal de Saúde do Rio de Janeiro e Clínica da Família Rinaldo Delamare, Rocinha, Rio de Janeiro, Brazil:** Solange Cavalcante, Betina Durovni, Jamile Garcia, Martinelle Godinho, João Marine

**Centro Municipal de Saúde de Duque de Caxias and Universidade Federal do Rio de Janeiro, Rio de Janeiro, Brazil:** Afranio Kritski, Adriana Moreira, André Luiz Bezerra, Anna Cristina Carvalho, Elisangela Silva

**Fundação de Medicina Tropical, Manaus, Brazil**: Marcelo Cordeiro dos Santos, Alexandra Brito, Leandro Garcia, Renata Cordeiro dos Santos, Allyson Souza, Jaquelane Silva

**Instituto Brasileiro para Investigação da Tuberculose, Fundação José Silveira, Salvador, Brazil:** Bruno de Bezerril Andrade, Eduardo M. Netto, Michael Rocha, Vanessa Nascimento, Betânia Nogueira, Saulo R. N. Santos, André Ramos

**Laboratório de Inflamação e Biomarcadores, Instituto Gonçalo Moniz, Fundação Oswaldo Cruz, Salvador, Brazil:** Alice Andrade, Hayna Malta, Juan Cubillos, Laise de Moraes

**Multinational Organization Network Sponsoring Translational and Epidemiological Research (MONSTER) Initiative, Salvador, Brazil:** Bruno de Bezerril Andrade, Artur Trancoso, Kiyosh Fukutani, Mariana Araujo Pereira, Maria Arriaga, Elze Leite

**Vanderbilt University Medical Center:** Gustavo Amorim, Timothy R. Sterling, David W. Haas, Marina C. Figueiredo, Megan Turner, Cody Staats, Brian Hachey

