## Supplemental Figures, Tables, and Appendix for "Estimating optimal therapeutic drug levels of anti-tuberculosis medications based on treatment safety and effectiveness"

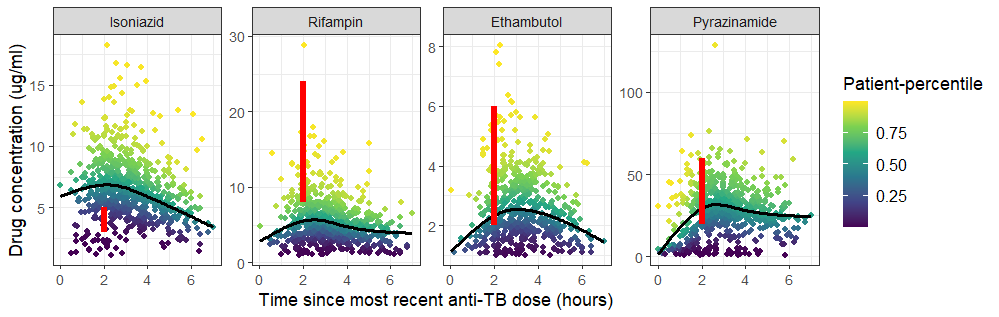
**Figure 1.** Drug concentration for each of the main four anti-TB drugs, colored by patient-percentiles. The red bars correspond to the currently recommended drug ranges.

**Table 1:** Summary data of all patients included in the study, stratified by their outcome status.

|  | [ALL]  (*N=448)* | No event  (*N=424)* | Grade 3 or higher ADR  (*N=9)* | Failure or Recurrence  *(N=15)* |
| --- | --- | --- | --- | --- |
| Age (in years) | 35.0 [25.0;47.0] | 35.0 [25.0;47.0] | 46.0 [38.0;60.0] | 38.0 [28.0;44.5] |
| Sex: Male | 304 (67.9%) | 290 (68.4%) | 5 (55.6%) | 9 (60.0%) |
| BMI | 19.9 [18.3;22.4] | 19.9 [18.3;22.4] | 21.2 [19.1;22.4] | 20.3 [17.6;22.0] |
| HIV status: Positive | 110 (24.6%) | 97 (22.9%) | 6 (66.7%) | 7 (46.7%) |
| Smoking status |  |  |  |  |
| Current | 95 (21.2%) | 88 (20.8%) | 3 (33.3%) | 4 (26.7%) |
| never | 218 (48.7%) | 207 (48.8%) | 6 (66.7%) | 5 (33.3%) |
| former | 135 (30.1%) | 129 (30.4%) | 0 (0.00%) | 6 (40.0%) |
| Alcohol status |  |  |  |  |
| current | 202 (45.1%) | 193 (45.5%) | 5 (55.6%) | 4 (26.7%) |
| never | 80 (17.9%) | 77 (18.2%) | 2 (22.2%) | 1 (6.67%) |
| former | 166 (37.1%) | 154 (36.3%) | 2 (22.2%) | 10 (66.7%) |
| Illicit drug use |  |  |  |  |
| current | 58 (12.9%) | 53 (12.5%) | 1 (11.1%) | 4 (26.7%) |
| never | 282 (62.9%) | 269 (63.4%) | 7 (77.8%) | 6 (40.0%) |
| former | 108 (24.1%) | 102 (24.1%) | 1 (11.1%) | 5 (33.3%) |
| Glycated hemoglobin (%) | 5.90 [5.50;6.40] | 5.90 [5.50;6.40] | 5.50 [5.30;5.70] | 6.40 [5.95;7.20] |
| AIMs proportion |  |  |  |  |
| African | 0.26 [0.14;0.43] | 0.26 [0.14;0.43] | 0.19 [0.13;0.37] | 0.21 [0.14;0.28] |
| European | 0.41 [0.28;0.53] | 0.41 [0.28;0.53] | 0.49 [0.33;0.68] | 0.42 [0.27;0.55] |
| Amerindian | 0.22 [0.14;0.38] | 0.22 [0.13;0.38] | 0.20 [0.11;0.28] | 0.34 [0.17;0.50] |
| DOT assigned: Yes | 21 (4.69%) | 18 (4.25%) | 0 (0.00%) | 3 (20.0%) |
| NAT2 acetylator: |  |  |  |  |
| rapid | 40 (8.93%) | 36 (8.49%) | 1 (11.1%) | 3 (20.0%) |
| intermediate | 174 (38.8%) | 165 (38.9%) | 5 (55.6%) | 4 (26.7%) |
| slow | 234 (52.2%) | 223 (52.6%) | 3 (33.3%) | 8 (53.3%) |

Note: Categorical variables are presented in terms of counts and percentages; continuous variables are presented as median and interquartile range.

Abbreviations: AIMs: Ancestry Informative Markers; DOT: direct observed therapy

**Table 2**: Log-odds for having an grade 3 or higher adverse drug reaction and the respective 95% confidence intervals. Estimates were obtained by fitting generalized estimating equations models with the logit link and an independent working correlation structure.

|  | Patient-percentile | | Cmax | |
| --- | --- | --- | --- | --- |
|  | **Log-odds (95% CI)** | **p-value** | **Log-odds (95% CI)** | **p-value** |
| INH | 0.53 (0.29; 0.77) | <0.01 | 5.18 (2.26; 8.10) | <0.01 |
| RIF | 0.76 (0.25; 1.27) | <0.01 | 4.38 (1.05; 7.71) | <0.01 |
| EMB | 0.22 (-0.05; 0.49) | 0.10 | 5.20 (0.16; 10.24) | 0.04 |
| PZA | 0.16 (-0.10; 0.41) | 0.22 | 1.29 (-0.66; 3.24) | 0.20 |

Abbreviations: CI: confidence interval; INH: Isoniazid; RIF: Rifampin; EMB: Ethambutol; PZA: Pyrazinamide.

**Table 3**: Log-odds for having failure/recurrence and the respective 95% confidence intervals. Estimates were obtained by fitting generalized estimating equations models with the logit link and an independent working correlation structure.

|  | Patient-percentile | | Cmax | |
| --- | --- | --- | --- | --- |
|  | **Log-odds (95% CI)** | **p-value** | **Log-odds (95% CI)** | **p-value** |
| INH | -0.16 (-0.29; -0.02) | 0.02 | -0.70 (-1.22; -0.17) | 0.01 |
| RIF | -0.08 (-0.20; 0.05) | 0.22 | -0.22 (-0.59; 0.14) | 0.23 |
| EMB | -0.05 (-0.22; 0.12) | 0.57 | -0.48 (-2.40; 1.44) | 0.62 |
| PZA | -0.02 (-0.19; 0.15) | 0.81 | 0.37 (-0.19; 0.93) | 0.19 |

Abbreviations: CI: confidence interval; INH: Isoniazid; RIF: Rifampin; EMB: Ethambutol; PZA: Pyrazinamide.

**Table 4**: Therapeutic drug ranges that are safe^1^ and effective^2^.

|  | Therapeutic drug range (µg/ml) | | |
| --- | --- | --- | --- |
|  | **Estimated range^3^** | | **Recommended^4^** |
|  | **Patient-percentile** | **Cmax** |  |
| INH | 1.62 (0; 2.30) - 11.00 (9.79; 13.30) | 2.31 (0; 3.88) - 8.22 (7.50; 11.00) | 3-5 |
| RIF | 0 (0; 0) - 9.07 (8.35; 10.02) | 0.34 (0; 1.58) - 7.49 (6.68; 9.71) | 8-24 |
| EMB | - | 0.17 (0; 0.82) - 3.01 (2.77; 3.98) | 2-6 |
| PZA | - | 0 (0; 2.40) - 49.5 (42.80; 65.90) | 20-50 |

Abbreviations: CI: confidence interval; INH: Isoniazid; RIF: Rifampin; EMB: Ethambutol; PZA: Pyrazinamide.

^1^ Probability of having adverse drug reaction lower than 5%

^2^ Probability of TB treatment failure/recurrence of at least 95%

^3^ Effectiveness bound (95% CI) - Safety bound (95% CI)

^4^ Peloquin CA. Therapeutic drug monitoring in the treatment of tuberculosis. Drugs 2002; 62: 2169–2183.

Dash symbols (-) denote scenarios in which the values could not be obtained.

**Appendix A.** Derivation of plasma drug exposures

We regressed the observed drug concentrations y against the timing t of the last TB dose (with a restricted cubic spline with 5 knots) to estimate the median of the drug concentration profiles via quantile regression. Quantile regression was used as an alternative to discretizing time intervals, reducing the subjective (if user-specified) or imprecise (when data is sparse) definition of the intervals. More discussion on the comparisons between the two approaches and the advantages of quantile regression for modeling population pharmacokinetic models can be found in Jamsem et. al. (2018).

The quantile regression estimates a set of parameters ***β*** such that

*min****_β_*** *Σ_ij_ |y_ij_ – f(t_ij_;* ***β****)|,*

where *f(t_ij_;* ***β****)* is a non-linear function on *t* indexed by ***β*** and *y_ij_* is the drug concentration from *j*th measurement collected from the *i*th individual at time *t_ij_*, for *i = 1, …, N*, and *j = 1, …,* *N_i_*, where *N* is the number of patients and *N_i_* is the number of blood samples collected for the *i*th person. The estimated parameters are then used to build a median drug concentration profile. This estimated profile indicates drug concentrations that are high or low for any given time *t_ij_*. That is, values above the median curve indicate drug concentrations that are higher than the expected median drug concentration, and values below suggest low drug concentrations.

We used the standardized difference between the observed and median expected drug *concentration* to generate the individual percentiles used in the analysis. More specifically, if we denote by *d_ij_* the difference between these two quantities and by *s* the standard deviation of the residuals obtained from the quantile regression, the individual standardized value *z_ij_* is equal to *d_ij_/s.* Notice that *z_ij_* indicates how many standard deviations the observed drug concentration *y_ij_* is from its estimated median value; values above the population median value are positive, while values below are negative.

The individual standardized values *z_ij_* were later mapped onto the interval [0,1] using the cumulative standard normal distribution function. Denote the mapped values as *p_ij_*. Although we could have used *z_ij_* in its natural scale, having *p_ij_*, varying from 0 to 1 is helpful for interpretation. For example, drug concentrations that were substantially higher than the expected median concentration will have a large *z_ij_* value and will, consequently, have large *p_ij_* (close to 1) or, equivalently, high percentages (100 x *p_ij_)*. Conversely, drug concentrations that were substantially lower than the median expected values will have a small *z_ij_* and, consequently, *p_ij_* and percentages values closer to 0. If both observed and median expected concentration coincided, *z_ij_* would be equal to 0, and *p_ij_* would be equal to 0.5 (50%).
